# Clinical aspects and serodiagnosis of myasthenia gravis in Algerian patients: a prospective study on a small group of patients

**DOI:** 10.1101/2025.01.19.25320522

**Authors:** Mohamed Nadji Bouchtout, William D Phillips, Elias Attal, Smail Daoudi, Sarah Kechoud, Stephen W Reddel, Chafia Touil-Boukoffa, Nabila Attal, Rachida Raache

## Abstract

Myasthenia gravis (MG) is a clinically heterogeneous disease, and its diagnosis relies primarily on serological tests such as ELISA and RIPA. This study reports the clinical characteristics of a cohort of Algerian patients with MG and evaluates the diagnostic accuracy of ELISA and RIPA for detecting anti-AChR and anti-MuSK autoantibodies. Serological analyses were performed using ELISA, RIPA, and cell-based assays (CBA) in a subset of 23 patients. Algerian MG patients did not exhibit any specific clinical features compared to other populations. The results demonstrated a good correlation between ELISA and RIPA for anti-AChR and anti-MuSK autoantibodies. However, 100% of patients with generalized MG and none of those with ocular MG were positive for anti-AChR by ELISA, compared to 83.3% and 40% by RIPA, respectively. None of the 23 patients were positive for anti-MuSK by RIPA or an in-house CBA. These findings suggest that anti-AChR ELISA may serve as a reliable diagnostic tool for MG in settings where RIPA is unavailable. Combining clinical and electrophysiological findings with serological testing may enhance the accuracy of MG diagnosis.

## 1. INTRODUCTION

Myasthenia gravis (MG) is an autoimmune disease caused by serum autoantibodies directed against components of the muscle membrane at the neuromuscular junction (NMJ). The disease is characterized by a failure of neuromuscular transmission that manifests as fluctuating muscle weakness (1). MG predominantly affects women aged between 20 and 39 and slightly more men than women aged between 50 and 70 (2). Most MG patients initially present with ocular symptoms (i.e., ptosis and/or diplopia). In about 75% of cases, other muscle groups are affected (i.e., bulbar, cervical, axial, proximal, distal, or respiratory muscle groups), typically within the first 2 to 3 years following presentation. When symptoms are solely ocular after 2 to 3 years following manifestation, MG is said to be ocular (OMG). When symptoms affect other muscle groups, MG is said to be generalized (GMG). The majority of GMG cases (80–85%) and half of OMG cases (50%) involve autoantibodies directed against the acetylcholine receptor (AChR), but a minority of GMG patients (5–10%) reveal autoantibodies directed against other NMJ components (1, 3–6), most commonly muscle-specific kinase (MuSK) (7, 8). MG pharmaceutical treatment relies primarily on acetylcholine inhibitors (e.g. pyridostigmine) and corticosteroids (e.g. prednisone) (9). Some MG patients present with a thymoma (10–20%), which requires surgical intervention, i.e., thymectomy (10) MG clinical severity is usually measured according to the Myasthenia Gravis Foundation of America (MGFA) classification (11).

The initial diagnosis of myasthenia gravis is usually based on clinical manifestation. Typically, this is followed by repetitive nerve stimulation electromyography (EMG). Radio immunoprecipitation assays (RIPA) are considered the gold standard assays for detecting AChR and MuSK autoantibodies. Enzyme-linked immunosorbent assays (ELISA) are less frequently used but are technically simpler and less costly, and these are important considerations in many parts of the world (12). Cell-based assays (CBA) are labor-intensive but are occasionally used as a last resort for MG patients when no autoantibody is detected by RIPA (13). Unlike RIPA and ELISA, CBAs detect the native antigen protein expressed on the surface of transfected mammalian cells, allowing them to sometimes detect cryptic autoantibodies (14). A small fraction of clinically diagnosed patients remain seronegative for both anti-AChR and anti-MuSK and might represent sensitivity limits of the assay or cases caused by antibodies against other synaptic antigens such as agrin and LRP4 (15).

Here we report the clinical characteristics of 67 Algerian MG patients and compare the anti-AChR and anti-MuSK status of 23 Algerian MG patients’ sera as assessed by ELISA and RIPA assays.

## 2. MATERIAL AND METHODS

### A. Clinical data and material

Sixty-seven Algerian patients with a high suspicion of MG were recruited for a clinical and serological assessment. Subsequently, twenty-three patients’ samples were selected for anti-MuSK RIPA, from which twelve were selected for anti-MuSK CBA. Sera from five congenital MG cases were used as controls. Patients were treated either at Ait Idir Neurosurgery Hospital (Algiers, Algeria) or at Sidi Belloua Hospital (Tizi Ouzou, Algeria). Informed patient consent was obtained from subjects in accordance with the Declaration of Helsinki. The study was approved by the University of Sciences and Technology Houari Boumediene (Algiers, Algeria) local ethics committee and the Algerian national agency of research development in health (ATRSS).

For each patient, the following information was recorded: disease age of onset (early onset [EOMG; ≤40 years], late onset [LOMG; between 41 and 65 years], and very late onset [VLOMG; ≥66 years]), sex (male or female), form (GMG or OMG), electromyographic profile (decrement ≥10% or <10%), current pharmaceutical treatment, thymus status (normal or presence of thymoma), whether the patient has undergone a thymectomy, and the MGFA score. All serum samples were collected within the 6 months following diagnosis and stored at +4°C. All patients were diagnosed with MG by their respective neurologists on the basis of clinical, electromyographic (EMG) and/or serological criteria.

### B. ELISA for anti-AChR

AChR antibody titres were assessed by an enzyme-linked immune-sorbent assay, following the manufacturers’ instructions (ELISA; ElisaRSR^TM^ AChRAb kit, RSR, Cardiff, UK) (16). This assay depends on the ability of human anti-AChR antibodies to compete for binding to the AChR with the two anti-AChR monoclonal antibodies provided in the kit, thereby inhibiting the ELISA signal. To summarize, 100 μl of patient serum was incubated overnight with 25 μl of a solubilized mixture of adult and foetal AChR. The sample-AChR mixture solution was then incubated in wells coated with anti-AChR antibodies. Wells were then washed and probed with biotinylated anti-AChR antibodies that were detected with Streptavidin-peroxydase and 3,3’, 5,5’-tetramethylbenzidine. Absorbance was read at 450 nm. The estimated human antibody concentration was expressed in nanomoles per liter of secondary IgG anti-human Fc. Only concentrations greater than 0.45 nmol/L were considered positive, as suggested by the manufacturer (RSR Ltd., Cardiff, UK).

### C. RIPA for anti-AChR and anti-MuSK

Commercially supplied ^125^I-AChR and ^125^I-MuSK, and RIPA kit including supplied standards were used, following the supplier’s protocol (RSR Ltd., Cardiff, UK) (17). Briefly, 5 µl of patient serum was incubated with 50 µl of ^125^I-AChR or ^125^I-MusK overnight at 4°C. Patient IgG was then immuno-precipitated by adding anti-human IgG, followed by centrifugation and a cold wash. The pelleted radioactivity was counted for 2 minutes on a gamma counter. Counts per minute were converted to nmol/L of secondary IgG anti-human Fc. Samples were considered negative for anti-AChR when values ranged from 0 to 0.25 nM, equivocal between 0.25 and 0.4 nM and positive above 0.4 nM. For anti-MuSK, 0 to 0.05 nM was considered negative, 0.05 to 0.09 nM equivocal and greater than 0.09 nM was considered positive.

### D. CBA for anti-MuSK

Published methods for the anti-MuSK CBA were followed (13, 18), with some modifications. Specifically, HEK293 cells were grown in Dulbecco’s modified eagle’s medium (DMEM) supplemented with 10% heat-inactivated fetal bovine serum (FBS), 1x antibiotic-antimycotic and 4 mM glutamine (Invitrogen™, MA, USA) at 37°C, 5% CO2. For transfection, 1.7×10^6^ cells were plated on a 60 mm culture dish. At 50-80% confluence, cells were transfected with 5 μg of expression plasmid encoding MuSK-EGFP (19) complexed with Lipofectamine LTX® Reagent (Invitrogen™, MA, USA). The next day, cells were replated onto 13 mm diameter glass coverslips in a 24-well tray (approximately 1×10^5^ cells/well). After a further 24 hours, wells were rinsed with DMEM-20 mM HEPES (pH 7.2) and incubated for 1 hour at room temperature with 300 µl of patient serum (1:20 dilution in 1% BSA/DMEM-20 mM HEPES). After 3 washes, coverslips were fixed with 3% paraformaldehyde, rinsed with phosphate-buffered saline (PBS), incubated for 45 mins with anti-human IgG-Texas Red (1:750; Invitrogen™, MA, USA), rinsed four times with PBS, permeabilized (5 mins in 0.02% Triton X100-PBS) and washed four times with PBS. Nuclei were counterstained with 4’,6-diamino-2-penylindole, dihydrochloride (DAPI; 2µg/ml in PBS). Coverslips were mounted on microscope slides using a fade-resistant mounting medium. A Zeiss Axio Imager fluorescence microscope and an AxioCamHRm digital camera were used to collect photomicrographs of 3 to 12 microscopic fields containing cells with peripheral membrane EGFP fluorescence for each sample. The exposure time was first optimized for the positive control coverslips (serum from two RIPA-positive Australian MG patients), and was held constant for all samples throughout the acquisition session. The CBA was performed twice (technical replicates). To avoid subjectivity, four volunteers were asked to blindly score all the digital photomicrographs. Blinding was achieved by using an online random number generator to rename each digital micrograph file. Volunteers were instructed to score the presence of anti-human IgG immunofluorescence co-localized on the surface of MuSK-EGFP-positive cells in each image as either definitely positive (score of 1), possibly positive (scores of 0.25, 0.5 or 0.75) or definitely negative (score of 0). Scores were converted into percentages. The criterion for considering a test sample positive by CBA was that it exceeded the mean + 5 SD of the values obtained for the non-myasthenic control.

### E. Statistical analysis

Means, standard deviations and/or percentages were calculated using the software GraphPad Prism 6. The paired t test was used to compare the means yielded by ELISA and RIPA. Pearson’s R was calculated to evaluate the linear correlation between the results of ELISA and RIPA. Histograms were generated using the software GraphPad Prism™ 8.0.1. Any *p* value lower than 0.05 (*p*<0.05) was considered statistically significant.

## 3. RESULTS

### A. Clinical characteristics

MG patients’ clinical characteristics are summarized in table 1. MG appeared before the age of 45 in 59.7% of patients, between the ages of 45 and 60 in 28.4% of patients, and after the age of 60 in 11.9% of patients. In male patients, MG developed after the age of 45 in 54.9% of patients, whereas in female patients, myasthenia developed in 72.2% of patients before the age of 45. Furthermore, 85.1% of patients had a GMG and only a fraction of 14.9% had OMG. Seropositivity for anti-AChR antibodies was greater than 91% in GMG but was equal to 20% in OMG.

**Table 1.**
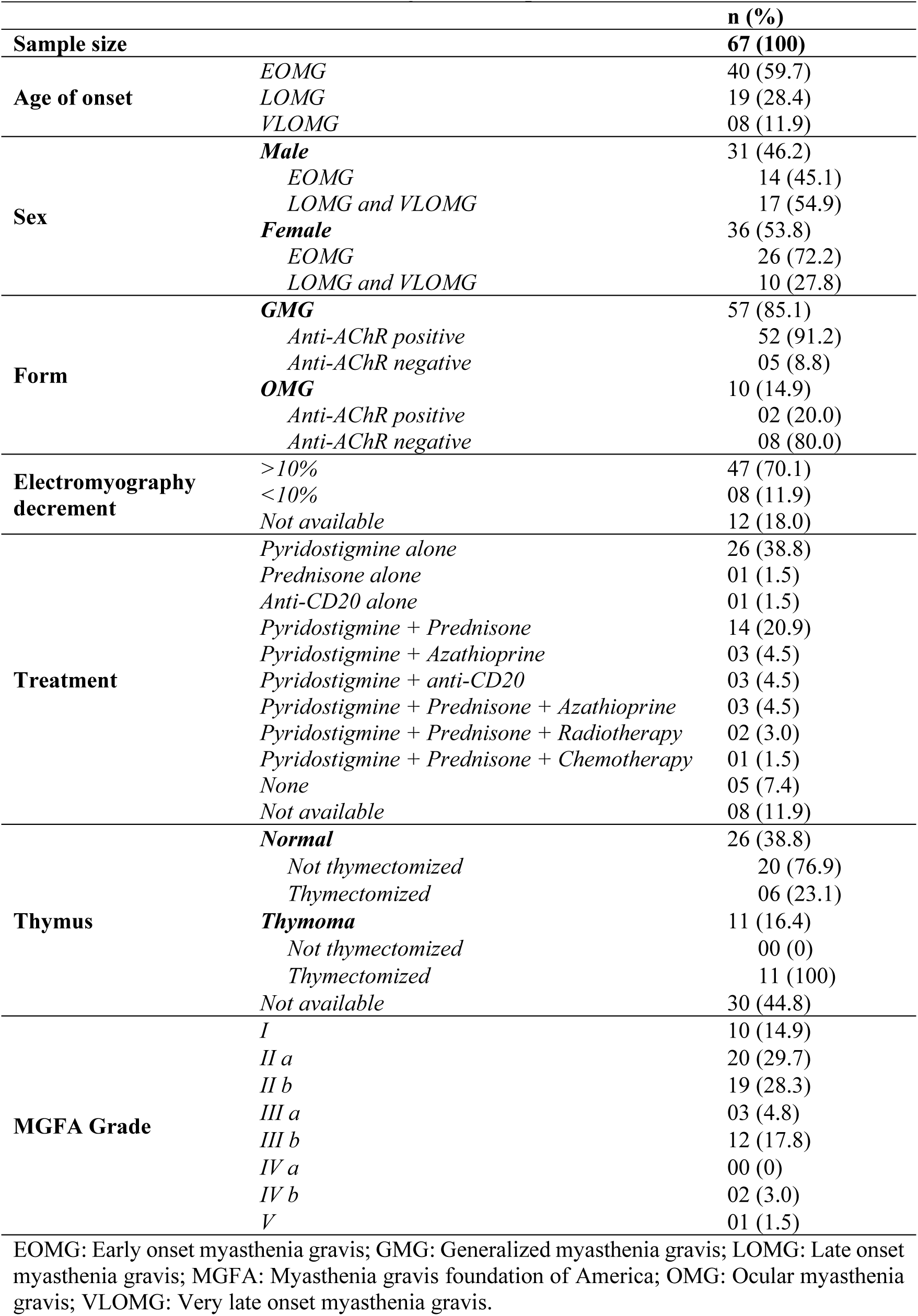
Clinical characteristics of Algerian MG patients.

|  |  | **n (%)** |
| --- | --- | --- |
| **Sample size** |  | **67 (100)** |
| **Age of onset** | *EOMG* | 40 (59.7) |
|  | *LOMG* | 19 (28.4) |
|  | *VLOMG* | 08 (11.9) |
| **Sex** | **Male** | 31 (46.2) |
|  | *EOMG* | 14 (45.1) |
|  | *LOMG and VLOMG* | 17 (54.9) |
|  | **Female** | 36 (53.8) |
|  | *EOMG* | 26 (72.2) |
|  | *LOMG and VLOMG* | 10 (27.8) |
| **Form** | **GMG** | 57 (85.1) |
|  | *Anti-AChR positive* | 52 (91.2) |
|  | *Anti-AChR negative* | 05 (8.8) |
|  | **OMG** | 10 (14.9) |
|  | *Anti-AChR positive* | 02 (20.0) |
|  | *Anti-AChR negative* | 08 (80.0) |
| **Electromyography decrement** | *>10%* | 47 (70.1) |
|  | *<10%* | 08 (11.9) |
|  | *Not available* | 12 (18.0) |
| **Treatment** | *Pyridostigmine alone* | 26 (38.8) |
|  | *Prednisone alone* | 01 (1.5) |
|  | *Anti-CD20 alone* | 01 (1.5) |
|  | *Pyridostigmine + Prednisone* | 14 (20.9) |
|  | *Pyridostigmine + Azathioprine* | 03 (4.5) |
|  | *Pyridostigmine + anti-CD20* | 03 (4.5) |
|  | *Pyridostigmine + Prednisone + Azathioprine* | 03 (4.5) |
|  | *Pyridostigmine + Prednisone + Radiotherapy* | 02 (3.0) |
|  | *Pyridostigmine + Prednisone + Chemotherapy* | 01 (1.5) |
|  | *None* | 05 (7.4) |
|  | *Not available* | 08 (11.9) |
| **Thymus** | **Normal** | 26 (38.8) |
|  | *Not thymectomized* | 20 (76.9) |
|  | *Thymectomized* | 06 (23.1) |
|  | **Thymoma** | 11 (16.4) |
|  | *Not thymectomized* | 00 (0) |
|  | *Thymectomized* | 11 (100) |
|  | *Not available* | 30 (44.8) |
| **MGFA Grade** | *I* | 10 (14.9) |
|  | *II a* | 20 (29.7) |
|  | *II b* | 19 (28.3) |
|  | *III a* | 03 (4.8) |
|  | *III b* | 12 (17.8) |
|  | *IV a* | 00 (0) |
|  | *IV b* | 02 (3.0) |
|  | *V* | 01 (1.5) |

EOMG: Early onset myasthenia gravis; GMG: Generalized myasthenia gravis; LOMG: Late onset myasthenia gravis; MGFA: Myasthenia gravis foundation of America; OMG: Ocular myasthenia gravis; VLOMG: Very late onset myasthenia gravis.

Among the 67 patients, 55 patients performed an EMG of which 47 (70.1%) showed a decrement greater than 10%, characteristic of MG. At the time of recruitment, 38.8% of the patients were on pyridostigmine alone and 20.9% were on a combination of pyridostigmine/prednisone. Other combinaisons (pyridostigmine/azathioprine, pyridostigmine/azathioprine/prednisone and pyridostigmine/anti-CD20) were less common (4.5%). Two patients (3%) were undergoing radiotherapy in addition to pyridostigmine/prednisone and one patient (1.5%) was undergoing chemotherapy in addition to pyridostigmine/prednisone. One patient (1.5%) was on prednisone alone while another patient was on anti-CD20 alone (1.5%). Lastly, a fraction of patients (7.4%) were not yet under any treatment.

Thoracic tomodensitometry was performed for 37 patients (55%) and showed that 38.8% of patients had a thymus without any abnormality whereas 16.4% had thymomas. Additionally, thymectomy was prescribed for 23.1% of patients with normal thymi and for all (100%) patients with thymomas.

Most patients (58%) had a mild form of MG that affected axial and limbs muscles as well as those of the bulbar region (MGFA grade IIa and IIb). In addition, 17.8% of patients had a moderate form, primarly bulbar (MGFA grade IIIb). A lesser proportion of patients (14.9%) had a strictly ocular form (grade MGFA I). Furthermore, 3% of patients had a severe form (MGFA grade IVb), characterized by a bulbar fatigability. Only one patient was admitted to the emergency department, was intubated and required respiratory assistance (MGFA grade V).

### B. Serological analysis

A subgroup of 23 patients (18 GMG and 5 OMG) were randomly selected for the serological analysis. Anti-AChR and anti-MuSK antibodies concentrations estimated by ELISA and RIPA and anti-MuSK seropositivity scores estimated by CBA, are presented in table 2.

**Table 2.** Anti-AChR and anti-MuSK antibodies concentrations or scores by ELISA, RIPA and/or CBA.

| Samples | Form | AChR ELISA<br>(nM) | AChR RIPA<br>(nM) | MuSK RIPA<br>(nM) | MuSK CBA ($M \pm \sigma$ %) | |
| --- | --- | --- | --- | --- | --- | --- |
|  |  |  |  |  | 1<sup>st</sup> run | 2<sup>nd</sup> run |
| 1 | GMG | 15.68 | 18.40 | 00.00 | 00 | 04.1 ± 08.3 |
| 2 | GMG | $\geq 20$ | 13.18 | 00.00 | 00 | 00 |
| 3 | GMG | 08.90 | 10.86 | 00.01 | 00 | 08.3 ± 06.7 |
| 4 | GMG | 13.69 | 09.35 | 00.00 | n/a | n/a |
| 5 | GMG | 08.20 | 04.86 | 00.00 | n/a | n/a |
| 6 | GMG | 07.82 | 08.10 | 00.01 | n/a | n/a |
| 7 | GMG | 18.35 | 19.31 | 00.00 | n/a | n/a |
| 8 | GMG | 16.42 | 03.79 | 00.00 | n/a | n/a |
| 9 | GMG | $\geq 20$ | 02.16 | 00.00 | n/a | n/a |
| 10 | GMG | 13.34 | 03.88 | 00.00 | n/a | n/a |
| 11 | GMG | 11.94 | 03.96 | 00.00 | n/a | n/a |
| 12 | GMG | 05.20 | 01.24 | 00.00 | 06.2 ± 07.9 | 00 |
| 13 | GMG | 14.51 | 02.97 | 00.01 | n/a | n/a |
| 14 | GMG | 05.13 | 00.01 | 00.00 | 20.8 ± 24.0 | 04.1 ± 04.7 |
| 15 | GMG | 04.90 | 00.12 | 00.01 | 02.0 ± 04.1 | 00 |
| 16 | OMG | 00.37 | 00.98 | 00.00 | 12.4 ± 10.7 | 06.2 ± 04.1 |
| 17 | OMG | 00.34 | 00.76 | 00.00 | 06.2 ± 04.1 | 00 |
| 18 | OMG | 00.44 | 00.15 | 00.00 | 08.3 ± 11.7 | 16.6 ± 23.5 |
| 19 | OMG | 00.38 | 00.21 | 00.00 | 00 | 00 |
| 20 | OMG | 00.41 | 00.12 | 00.01 | 02.0 ± 04.1 | 00 |
| 21 | GMG | 15.57 | 16.42 | 00.00 | n/a | n/a |
| 22 | GMG | 00.25 | 00.05 | 00.00 | 24.9 ± 16.6 | 00 |
| 23 | GMG | 00.65 | 00.26 | 00.01 | n/a | n/a |
| CMS 1 | / | 00.28 | 00.00 | 00.01 | 14.5 ± 14.2 | 00 |
| CMS 2 | / | 00.42 | 00.14 | 00.01 | 12.5 ± 14.4 | 00 |
| CMS 3 | / | 00.25 | 00.15 | 00.00 | 00 | 02.0 ± 04.1 |
| CMS 4 | / | 00.25 | 00.14 | 00.00 | 00 | 00 |
| CMS 5 | / | 00.30 | 00.12 | 00.00 | 00 | 04.1 ± 04.7 |
| MuSKPC1 | n/a | n/a | 00.15 | 00.84 | 95.8 ± 8.3 | 100 |
| MuSKPC2 | n/a | n/a | 00.13 | 00.83 | 100 | 100 |
| MuSKNC1 | / | n/a | 00.39 | 00.01 | 00 | 02.0 ± 04.1 |
| MuSKNC2 | / | n/a | 00.19 | 00.01 | 04.1 ± 04.7 | 00 |
| Threshold | / | 00.45 | 00.40 | 00.09 | 27.6 | 22.5 |

AChR : Acetylcholine receptor ; CBA : Cell-based assay ; CMS: Congenital myasthenic syndrome ; ELISA : Enzyme-linked immunosorbent assay ; $M \pm \sigma$ %: Mean ± standard deviation in percentages ; MuSK : Muscle-specific kinase ; MuSKNC : Anti-MuSK negative control ; MuSKPC : Anti-MuSK positive control ; n/a : Not available ; nM : Nanomoles per liter.

Anti-AChR ELISA and anti-AChR RIPA results comparison and correlation are illustrated in Fig. 1. The mean concentration (± standard deviation) estimated by ELISA was 8.8 ± 7.1 nM and that estimated by RIPA was 5.2 ± 6.3 nM. These two mean concentrations were significantly different (*p*=0.0045; Fig. 1A). Nevertheless, our results showed the existence of a moderate positive correlation between the results of the two assays (slope: 0.60; Pearson’s R^2^=0.47; p=0.0003; Fig. 1B) even though some patients results may differ substantially (e.g., patients 8 to 11, table 2). In addition, all GMG patients (100%) were seropositive by ELISA whereas 83.3% were RIPA positive. Conversely, not a single OMG patient (0%) was seropositive by ELISA whereas 40% were RIPA positive (table 3).

**Figure 1:**
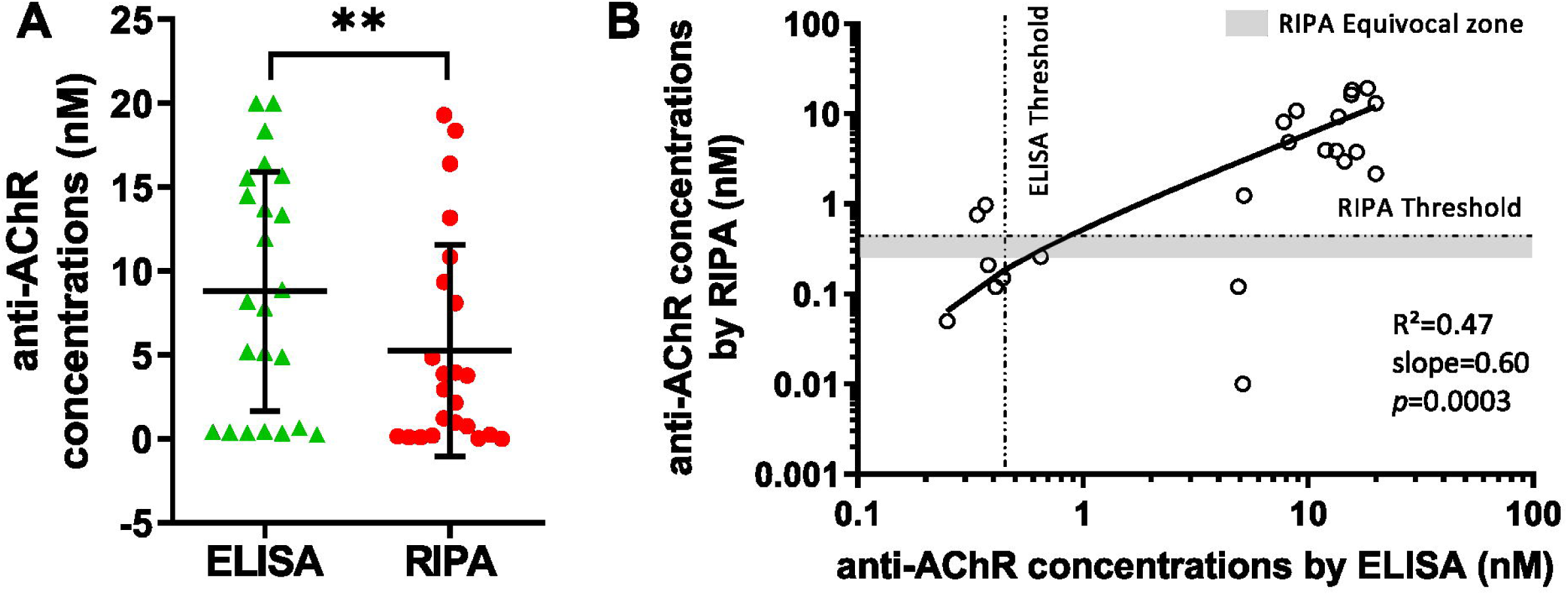
Anti-AChR antibodies results comparison. (A) Comparison of anti-AChR concentrations obtained by ELISA with those obtained by RIPA. (B) Correlation between anti-AChR concentrations obtained by ELISA versus those obtained by RIPA. Each symbol represents the results for an individual MG patient.

**Table 3.**
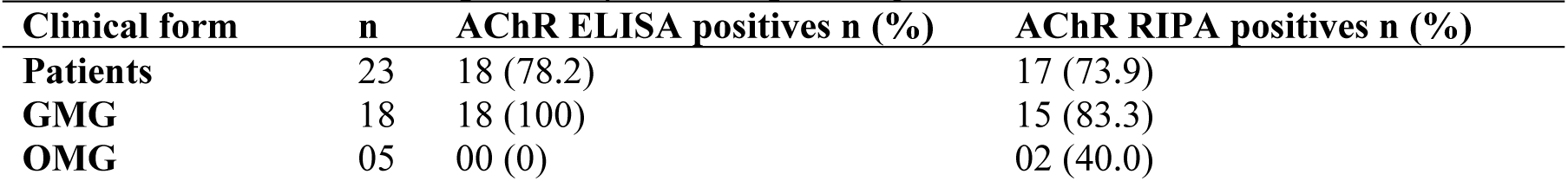
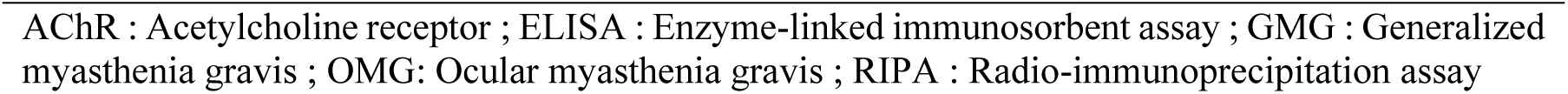
Anti-AChR seropositivity rates depending on the clinical form.

| Clinical form | n | AChR ELISA positives n (%) | AChR RIPA positives n (%) |
| --- | --- | --- | --- |
| Patients | 23 | 18 (78.2) | 17 (73.9) |
| GMG | 18 | 18 (100) | 15 (83.3) |
| OMG | 05 | 00 (0) | 02 (40.0) |

AChR : Acetylcholine receptor ; ELISA : Enzyme-linked immunosorbent assay ; GMG : Generalized myasthenia gravis ; OMG: Ocular myasthenia gravis ; RIPA : Radio-immunoprecipitation assay

None of the 23 Algerian MG patients’ sera, nor the 5 Algerian congenital myasthenic syndrome patients’ sera were positive for anti-MuSK by RIPA (table 1). We rescreened a subset of 12 of the Algerian MG samples using a CBA (see Methods). Fig. 2A shows an example of cell surface immunofluorescence with an anti-MuSK positive serum (by RIPA) from an Australian MG patient (Fig. 2A: human IgG immunofluorescence decorating the periphery of the MuSK-EGFP-expressing cell). Negative control serum showed no such IgG immunolabeling (Fig. 2B). Four volunteers blindly scored images for cell surface IgG labeling that co-localized with MuSK-GFP, after first being trained with positive and negative control micrographs. The CBA was run twice with the same samples, so as to provide technical replication. All four raters correctly scored images from the anti-MuSK positive-controls sera (first run = 95.8 ± 8.3% and 100% [mean ± SD]; replicate run = 100 ± 0% for both controls). Negative-control images were also correctly identified (first run = 2.0 ± 4.1% and 0%; replicate run = 4.1 ± 4.7% and 0%). Thus, all 4 blinded scorers were able to clearly distinguish positive and negative control samples. When assessing the CBA images for the 12 Algerian samples and even though the 4 blinded scorers revealed some variability, no mean score was found unambiguously greater than the positivity threshold in both replicates (Fig. 2C and D). In summary, neither RIPA nor CBA revealed anti-MuSK positive MG patients among the Algerian cohort tested.

**Figure 2:**
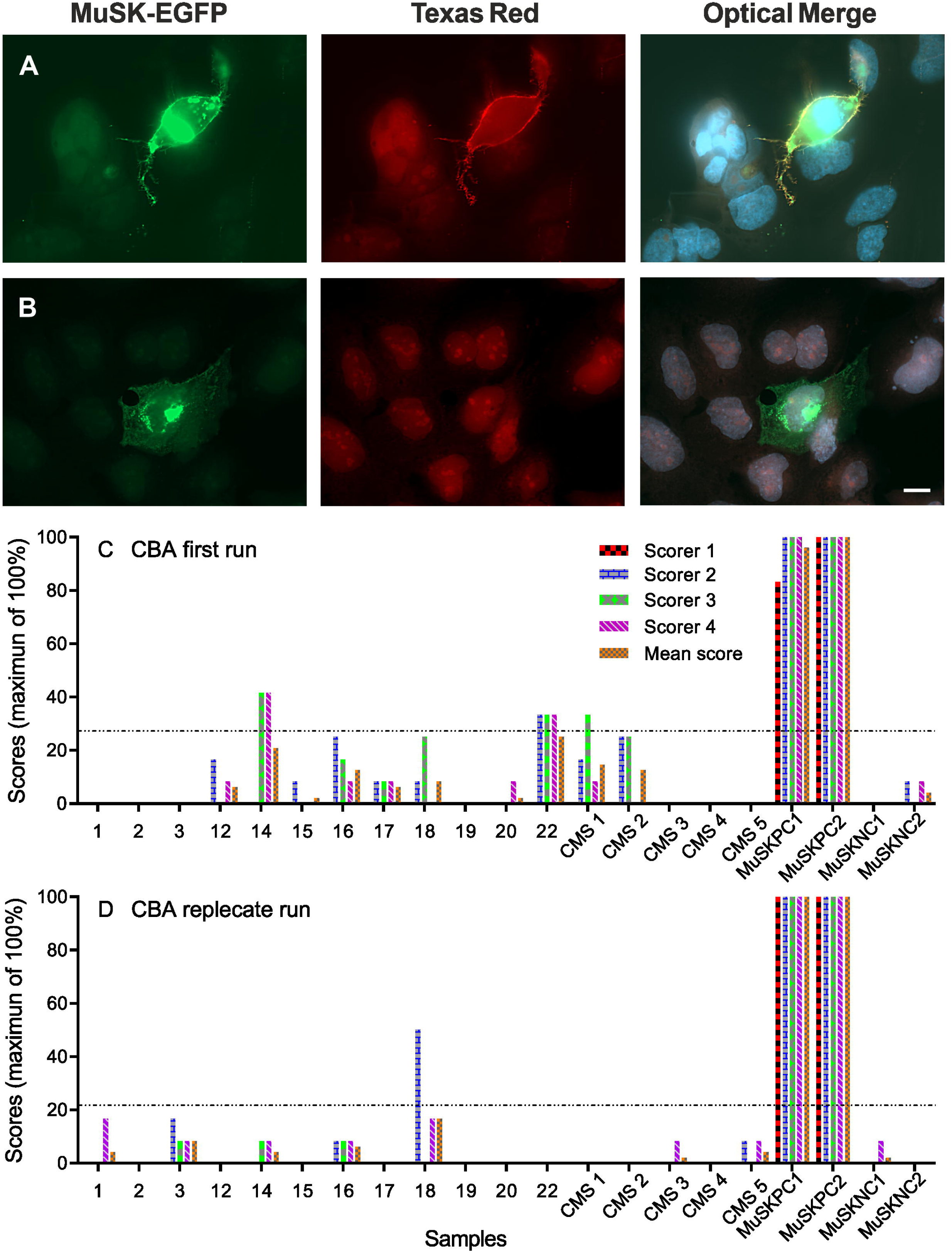
MuSK CBA results. (A) Immunofluorescence reveals human antibody labeling co-localizing with MuSK-EGFP on the surface of transfected HEK293 cells. Live cells were exposed to anti-MuSK-positive control serum and human IgG was detected with Texas red conjugated secondary antibody. Only a fraction of these transiently transfected cells expressed MuSK-EGFP, the surrounding (non-expressing) cells provide an indication of the level of non-specific background fluorescence. (B) Cultures incubated with RIPA-negative control serum showed no such co-localization of human IgG with cell surface MuSK-EGFP (scale bar = 4 μm). (C & D) Comparison of results from duplicate CBAs. Four blinded observers each independently scored colocalization of Texas red immunofluorescence with MuSK-EGFP on cells. Bars at far right show results for positive control and negative control sera respectively. CMS 1 to 5 show results of sera from patients with myasthenic congenital syndrome. The remaining show results for Algerian MG patients. Horizontal dashed lines indicate a threshold of mean + 5 SD of the negative control sample scores. CMS: congenital myasthenic syndrome; MuSKPC: positive control; MuSKNC: negative control.

## 4. DISCUSSION

Various clinical parameters of 67 Algerian patients with myasthenia gravis were reported. Our results show that MG affects mostly females under the age of 45, which is in agreement with different previous studies conducted on Japanese, Norwegian and Portuguese populations (20–22). However, some studies suggest a predominance of incidence in individuals above the age of 65 years in European and Australian populations (23–26). The incidence of MG in the elderly is reportedly increasing in Western countries, Australia, as well as in Japan. Our results do not indicate such an increase in the Algerian population. This difference in incidence could be due to genetic and/or demographic differences (27). This is all the more plausible considering that the proportions of seniors in the Western and Japanese populations, compared to the Algerian population, differ considerably (percentage of individuals over 65 years old in 2019: 15.9, 18, 28 and 6.6% for Australia, Europe, Japan and Algeria, respectively) (28). Moreover, the difference in the mean age at disease onset of the patients included in our study (40 years) and those of other studies (59 years in an Australian epidemiological study (24) and 58 years in a Spanish multicenter study (25) for example) also seems considerable. Thus, the relatively low proportion of individuals over 65 years of age in Algeria could explain the absence of the new incidence peak.

We also report that GMG affects 85% of patients (vs. 15% OMG) and that seropositivity for anti-AChR is far greater in GMG than in OMG, which corroborates with what has been consistently reported (4, 29). Nevertheless, anti-AChR antibodies have been detected in only 20% of our OMG patients, which is below the usually reported proportion of approximately 50% (30). This low proportion might reflect a lack of sensitivity in the anti-AChR ELISA technique.

EMG is considered an important first-line examination to demonstrate a failure of neuromuscular transmission. It is reported that 30 to 80% of MG patients have an abnormal EMG and our results (70%) are in agreement with those reported (31). Furthermore, our results seem to reflect good adherence to the International Consensus Guidelines for the Management of MG (9) since about 40% of the patients were on pyridostigmine monotherapy and about 20% of the patients were on dual therapy with pyridostigmine and prednisolone. In addition, about 15% of the patients were on dual therapy with azathioprine or anti-CD20 in combination with pyridostigmine or on triple therapy with prednisolone. Thus, it would seem that the first, second- and third-line treatments as recommended by international standards are suitable for Algerian patients. Moreover, 16% of the patients included in our study had a thymoma, which is consistent with what has been reported in the literature (32).

MG presents particular difficulties for the “measurement” of the patient’s condition since the classical manifestation of MG (fatigability) is translated by signs and symptoms that fluctuate in time and with effort. Measurement of MG symptoms according to the MGFA classification is the most commonly used method and aims to classify cases into groups based on disease severity and symptom location (11). In our study, the majority (58%) of patients had a mild GMG (groupe II), and our results are in agreement with those reported by various studies conducted on Italian, Brazilian, Japanese, and Austrian populations, where the proportion of patients in group II varied between 45 and 70% (33–36).

In this study, 23 Algerian myasthenia gravis patients’ sera samples were tested for anti-AChR by ELISA and RIPA. Both assays yielded comparable results for most patient samples. Estimates of serum anti-AChR concentration by ELISA and RIPA showed a clear correlation. Nevertheless, the average anti-AChR concentration by ELISA was higher than RIPA, which suggests that ELISA tends to overestimate anti-AChR concentration due to a lower specificity than RIPA. Taken separately, our anti-AChR RIPA results were comparable to those reported in various reviews (anti-AChR GMG: 83.3%; anti-AChR OMG: 40%) (1, 4, 14) and show that Algerian MG patients’ seropositivity ratio is not obviously different from that reported in other populations. However, anti-AChR ELISA results were somewhat different (GMG: 100% positive; OMG: 0%). Anti-AChR ELISA detected antibodies in three GMG patients (16.7%) who tested negative by anti-AChR RIPA. These results are in line with those reported by Oger & Frykman (12) where they re-screened 40 healthy individuals’ samples (negative by RIPA) with the Euroimmune™ ELISA and detected 2 (5%) false positives. Their results and ours suggest that some false positives can be expected by anti-AChR ELISA. However, it remains possible that “false positive” samples might contain genuine AChR autoantibody that happens to bind more efficiently when the antigen is presented in a solid phase ELISA assay, compared to the liquid phase RIPA assay. Retesting such samples in a well-validated CBA might help resolve this issue. Conversely, among the OMG samples, two patients that were positive by anti-AChR RIPA were found to be negative by anti-AChR ELISA. The results yielded by anti-AChR RIPA for these samples were slightly above the recommended threshold whereas anti-AChR ELISA results were below the suggested threshold, consistent with the idea that ELISA may be less likely to detect low concentration/low affinity anti-AChR when compared to testing by anti-AChR RIPA (37). Our results corroborate once more with those of Oger & Frykman (12) who reported that out of 90 samples that were anti-AChR positive by RIPA, only 66 were positive by ELISA (27% false negatives). Furthermore, our results suggest that ELISA false seropositivity is more likely to be observed in GMG due to a relatively low specificity, whereas false seronegativity is mainly recorded in OMG due to a relatively low sensitivity. However, a study compared a commercial ELISA, an in-house ELISA, and an in-house Dot Blot for the detection of anti-AChR antibodies in MG patients’ sera and reported that 92.2% of GMG patients and 52.9% of OMG patients were positive in all three assays (38). The same study reported that no false positives were found after screening 85 samples from healthy individuals by all three assays. Our results are not in line with those reported by Bokoliya et al. (2019) and this might be due to technical differences related to the commercial kits (ElisaRSR™ AChRAb *vs.* Novus Biologicals™ Acetylcholine ELISA) and/or to the small number of patients included in our study.

The percentage of MG patients reported to express autoantibodies targeting MuSK rather than AChR varies widely (10% to 70% in anti-AChR seronegative patients) (39–41) In the present study, the anti-MuSK RIPA assay revealed no anti-MuSK-positive serum samples among the 23 Algerian MG patients. Similarly, none of the 12 patients’ serum samples were positive when re-tested by CBA despite the strong labeling observed with serum from two Australian anti-MuSK seropositive control patients. If a CBA result had been based solely upon a single scorer in a single run, several false-positives might have resulted. Nevertheless, when the results from several scorers were averaged, the scores for all the Algerian patients were comparable to those for the Australian negative-control patient samples. Although we did not assess our CBA specificity and sensitivity due to a lack of anti-MuSK positive samples, we were nevertheless able to establish a CBA that has acceptable reliability and a scoring system that allowed correct interpretation of the results. Cell-based assays are increasingly studied in the context of MG serodiagnosis and seem to be very promising. (14, 15, 42–46)

## 5. CONCLUSION

In summary, the present study showed that Algerian MG patients present typical clinical and serological profiles. We also show that in a cost-constrained health care system, the anti-AChR ELISA kit used in this study can be applied for diagnostic purposes with a reasonable level of accuracy. Our findings also suggest that some results from anti-AChR ELISA need to be interpreted with caution. False positives and false negatives were previously reported to be non-negligible and our results support this concern. While RIPA remains the gold standard assay for anti-AChR antibodies detection, many diagnostic laboratories around the world lack facilities for handling and assaying radioisotopes. In this study, we did not have sufficient anti-AChR seronegative cases to properly explore for anti-MuSK antibodies.

We suggest that, in cost-constrained health systems, ELISA can be used as the primary screening test. Retesting or sending away samples for secondary analysis by RIPA and/or CBA might be limited to cases of diagnostic doubt or for cases of acquired MG that are well characterized by clinical and electrophysiological criteria yet seronegative by ELISA. The availability of commercial anti-AChR ELISA kits and the relative simplicity of its realization make the anti-AChR ELISA assay useful when used in combination with clinical observations and EMG for the diagnosis of MG.

## Data Availability

All data produced in the present study are available upon reasonable request to the authors

## REFERENCES

1. Berrih-Aknin S, Le Panse R. Myasthenia gravis: a comprehensive review of immune dysregulation and etiological mechanisms. J Autoimmun. 2014;52:90–100. doi: 10.1016/j.jaut.2013.12.011.

2. Bubuioc A-M, Kudebayeva A, Turuspekova S, Lisnic V, Leone MA. The epidemiology of myasthenia gravis. J Med Life. 2021;14(1):7. doi: 10.25122/jml-2020-0145.

3. Lindstrom JM, Seybold ME, Lennon VA, Whittingham S, Duane DD. Antibody to acetylcholine receptor in myasthenia gravis: prevalence, clinical correlates, and diagnostic value. Neurology. 1976;26(11):1054-. doi: 10.1212/WNL.51.4.933-a.

4. Phillips WD, Vincent A. Pathogenesis of myasthenia gravis: update on disease types, models, and mechanisms. F1000Research. 2016;5. doi: 10.12688/f1000research.8206.1.

5. Vincent A. Unravelling the pathogenesis of myasthenia gravis. Nature Reviews Immunology. 2002;2(10):797–804. doi: 10.1038/nri916.

6. Ha JC, Richman DP. Myasthenia gravis and related disorders: Pathology and molecular pathogenesis. Biochimica et Biophysica Acta (BBA)-Molecular Basis of Disease. 2015;1852(4):651–7. doi: 10.1016/j.bbadis.2014.11.022.

7. Hoch W, McConville J, Helms S, Newsom-Davis J, Melms A, Vincent A. Auto-antibodies to the receptor tyrosine kinase MuSK in patients with myasthenia gravis without acetylcholine receptor antibodies. Nat Med. 2001;7(3):365–8. doi: 10.1038/85520.

8. Koneczny I, Cossins J, Vincent A. The role of muscle-specific tyrosine kinase (MuSK) and mystery of MuSK myasthenia gravis. J Anat. 2014;224(1):29–35. doi: 10.1111/joa.12034.

9. Narayanaswami P, Sanders DB, Wolfe G, Benatar M, Cea G, Evoli A, et al. International consensus guidance for management of myasthenia gravis: 2020 update. Neurology. 2021;96(3):114–22. doi: 10.1212/WNL.000000000001112.

10. Frykman H, Kumar P, Oger J. Immunopathology of Autoimmune Myasthenia Gravis: Implications for Improved Testing Algorithms and Treatment Strategies. Front Neurol. 2020;11:1685. doi: 10.3389/fneur.2020.596621.

11. Barnett C, Herbelin L, Dimachkie MM, Barohn RJ. Measuring clinical treatment response in myasthenia gravis. Neurol Clin. 2018;36(2):339–53. doi: 10.1016/j.ncl.2018.01.006.

12. Oger J, Frykman H. An update on laboratory diagnosis in myasthenia gravis. Clin Chim Acta. 2015;449:43–8. doi: 10.1016/j.cca.2015.07.030.

13. Leite MI, Jacob S, Viegas S, Cossins J, Clover L, Morgan BP, et al. IgG1 antibodies to acetylcholine receptors in ‘seronegative’myasthenia gravis. Brain. 2008;131(7):1940–52. doi: 10.1093/brain/awn092.

14. Gastaldi M, Scaranzin S, Businaro P, Mobilia E, Benedetti L, Pesce G, et al. Improving laboratory diagnostics in myasthenia gravis. Expert review of molecular diagnostics. 2021;21(6):579–90. doi: 10.1080/14737159.2021.1927715.

15. Lazaridis K, Tzartos SJ. Autoantibody specificities in myasthenia gravis; implications for improved diagnostics and therapeutics. Front Immunol. 2020;11:212. doi: 10.3389/fimmu.2020.00212.

16. Hewer R, Matthews I, Chen S, McGrath V, Evans M, Roberts E, et al. A sensitive non-isotopic assay for acetylcholine receptor autoantibodies. Clin Chim Acta. 2006;364(1-2):159–66. doi: 10.1016/j.cccn.2005.05.035.

17. Matthews I, Chen S, Hewer R, McGrath V, Furmaniak J, Smith BR. Muscle-specific receptor tyrosine kinase autoantibodies—a new immunoprecipitation assay. Clin Chim Acta. 2004;348(1-2):95–9. doi: 10.1016/j.cccn.2004.05.008.

18. Huda S, Waters P, Woodhall M, Leite MI, Jacobson L, De Rosa A, et al. IgG-specific cell-based assay detects potentially pathogenic MuSK-Abs in seronegative MG. Neurology-Neuroimmunology Neuroinflammation. 2017;4(4). doi: 10.1212/NXI.000000000000035.

19. Cole R, Ghazanfari N, Ngo S, Gervasio O, Reddel S, Phillips W. Patient autoantibodies deplete postsynaptic muscle-specific kinase leading to disassembly of the ACh receptor scaffold and myasthenia gravis in mice. The Journal of physiology. 2010;588(17):3217–29. doi: 10.1113/jphysiol.2010.190298.

20. Murai H, Yamashita N, Watanabe M, Nomura Y, Motomura M, Yoshikawa H, et al. Characteristics of myasthenia gravis according to onset-age: Japanese nationwide survey. J Neurol Sci. 2011;305(1-2):97–102. doi: 10.1016/j.jns.2011.03.004.

21. Andersen J, Heldal A, Engeland A, Gilhus N. Myasthenia gravis epidemiology in a national cohort; combining multiple disease registries. Acta Neurol Scand. 2014;129:26–31. doi: 10.1111/ane.12233.

22. Santos E, Bettencourt A, da Silva AM, Boleixa D, Lopes D, Brás S, et al. HLA and age of onset in myasthenia gravis. Neuromuscul Disord. 2017;27(7):650–4. doi: 10.1016/j.nmd.2017.04.002.

23. McGrogan A, Sneddon S, De Vries CS. The incidence of myasthenia gravis: a systematic literature review. Neuroepidemiology. 2010;34(3):171–83. doi: 10.1159/000279334.

24. Gattellari M, Goumas C, Worthington J. A national epidemiological study of Myasthenia Gravis in Australia. Eur J Neurol. 2012;19(11):1413–20. doi: 10.1111/j.1468-1331.2012.03698.x.

25. Cortés-Vicente E, Álvarez-Velasco R, Segovia S, Paradas C, Casasnovas C, Guerrero-Sola A, et al. Clinical and therapeutic features of myasthenia gravis in adults based on age at onset. Neurology. 2020;94(11):e1171–e80. doi: 10.1212/WNL.0000000000008903.

26. Belimezi M, Kalliaropoulos A, Jiménez J, Garcia I, Mentis A-FA, Chrousos GP. Age at sampling and sex distribution of AChRAb vs. MuSKAb myasthenia gravis in a large Greek population. Clin Neurol Neurosurg. 2021;208:106847. doi: 10.1016/j.clineuro.2021.106847.

27. Carr AS, Cardwell CR, McCarron PO, McConville J. A systematic review of population based epidemiological studies in Myasthenia Gravis. BMC Neurol. 2010;10(1):1–9. doi: 10.1186/1471-2377-10-46.

28. United Nations, Department of Economic and Social Affairs, Population Division (2019). World Population Ageing 2019: Highlights (ST/ESA/SER. A/430).

29. Hendricks TM, Bhatti MT, Hodge DO, Chen JJ. Incidence, epidemiology, and transformation of ocular myasthenia gravis: a population-based study. Am J Ophthalmol. 2019;205:99–105. doi: 10.1016/j.ajo.2019.04.017.

30. Ciafaloni E. Myasthenia gravis and congenital myasthenic syndromes. Continuum: Lifelong Learning in Neurology. 2019;25(6):1767–84. doi:10.1212/CON.0000000000000800.

31. Anh Pham TK, De Tran V, Nguyen KT, Pham PV, Dewey RS, Nguyen BT, Le TD. Characterization of myasthenia gravis using clinical classification and repetitive nerve stimulation. Arch Balk Med Union. 2021;56(2):165–171. doi: 10.31688/ABMU.2021.56.2.04.

32. Álvarez-Velasco R, Gutiérrez-Gutiérrez G, Trujillo JC, Martínez E, Segovia S, Arribas-Velasco M, et al. Clinical characteristics and outcomes of thymoma-associated myasthenia gravis. Eur J Neurol. 2021;28(6):2083–91. doi: 10.1111/ene.14820.

33. Mantegazza R, Beghi E, Pareyson D, Antozzi C, Peluchetti D, Sghirlanzoni A, et al. A multicentre follow-up study of 1152 patients with myasthenia gravis in Italy. J Neurol. 1990;237(6):339–44. doi: 10.1007/BF00315656.

34. Werneck L, Cunha F, Scola R. Myasthenia gravis: a retrospective study comparing thymectomy to conservative treatment. Acta Neurol Scand. 2000;101(1):41–6. doi: 10.1034/j.1600-0404.2000.00014.x.

35. Kawaguchi N, Kuwabara S, Nemoto Y, Fukutake T, Satomura Y, Arimura K, et al. Treatment and outcome of myasthenia gravis: retrospective multi-center analysis of 470 Japanese patients, 1999–2000. J Neurol Sci. 2004;224(1-2):43–7. doi: 10.1016/j.jns.2003.09.016.

36. Rath J, Brunner I, Tomschik M, Zulehner G, Hilger E, Krenn M, et al. Frequency and clinical features of treatment-refractory myasthenia gravis. J Neurol. 2020;267(4):1004–11. doi: 10.1007/s00415-019-09667-5.

37. Swanson SJ, Ferbas J, Mayeux P, Casadevall N. Evaluation of methods to detect and characterize antibodies against recombinant human erythropoietin. Nephron Clinical Practice. 2004;96(3):c88–c95. doi: 10.1159/000076746.

38. Bokoliya S, Patil S, Nagappa M, Taly A. A simple, rapid and non-radiolabeled immune assay to detect anti-AChR antibodies in myasthenia gravis. Lab Med. 2019;50(3):229–35. doi: 10.1093/labmed/lmy038.

39. Berrih-Aknin S, Frenkian-Cuvelier M, Eymard B. Diagnostic and clinical classification of autoimmune myasthenia gravis. J Autoimmun. 2014;48:143–8. doi: 10.1016/j.jaut.2014.01.003.

40. Phillips W, Christadoss P, Losen M, Punga AR, Shigemoto K, Verschuuren J, et al. Guidelines for pre-clinical animal and cellular models of MuSK-myasthenia gravis. Exp Neurol. 2015;270:29–40. doi: 10.1016/j.expneurol.2014.12.013.

41. Evoli A, Alboini PE, Damato V, Iorio R, Provenzano C, Bartoccioni E, et al. Myasthenia gravis with antibodies to MuSK: an update. Ann N Y Acad Sci. 2018;1412(1):82–9. doi: 10.1016/j.expneurol.2014.12.013.

42. Mirian A, Nicolle MW, Edmond P, Budhram A. Comparison of fixed cell-based assay to radioimmunoprecipitation assay for acetylcholine receptor antibody detection in myasthenia gravis. J Neurol Sci. 2022;432:120084. doi: 10.1016/j.jns.2021.120084.

43. Han J, Zhang J, Li M, Zhang Y, Lv J, Zhao X, et al. A novel MuSK cell-based myasthenia gravis diagnostic assay. J Neuroimmunol. 2019;337:577076. doi: 10.1016/j.jneuroim.2019.577076.

44. Damato V, Spagni G, Monte G, Woodhall M, Jacobson L, Falso S, et al. Clinical value of cell-based assays in the characterisation of seronegative myasthenia gravis. J Neurol Neurosurg Psychiatry. 2022;93(9):995–1000. doi: 10.1136/jnnp-2022-329284.

45. Kim MJ, Kim SW, Kim M, Choi YC, Kim SM, Shin HY, et al. Evaluating an in-house cell-based assay for detecting antibodies against muscle-specific tyrosine kinase in myasthenia gravis. J Clin Neurol. 2021;17(3):400–408. doi: 10.3988/jcn.2021.17.3.400.

46. Spagni G, Gastaldi M, Businaro P, Chemkhi Z, Carrozza C, Mascagna G, et al. Comparison of fixed and live cell-based assay for the detection of AChR and MuSK antibodies in myasthenia gravis. Neurol Neuroimmunol Neuroinflamm. 2022;10(1):e200038. doi: 10.1212/NXI.0000000000200038.

